# Progress in Epidemic Ready Primary Health Care: Early Pilot Results from Four African Countries (Ethiopia, Nigeria, Sierra Leone and Uganda), December 2023 – October 2024

**DOI:** 10.1101/2025.02.14.25322294

**Authors:** Stacey Mearns, Moreen Kamateeka, Tochi Okwor, Aisha Abba, Celestina Obiekea, Jenom Danjuma, Arone M Haile, Daniel Damtew, Damene Debalke, Joseph S Kanu, Ramatu Ngauja, Susan Michaels-Strasser, Abdullah Wailagala, Allan Muruta, Atim Dansan, Amy Barrera-Cancedda, Samantha Kozikott, Justine Landegger, Leena Patel, Amanda McClelland

## Abstract

Primary healthcare (PHC) is the first point of contact with communities and essential for epidemic preparedness. The COVID-19 pandemic exposed gaps in PHC resilience. Epidemic Ready Primary Health Care (ERPHC) was designed to bridge these gaps by strengthening PHC to prevent, detect, and respond to outbreaks while maintaining essential services. An ERPHC pilot was initiated in December 2023 in 654 PHC facilities across Ethiopia, Nigeria, Sierra Leone, and Uganda. The approach improves connection to local communities, detection and reporting of cases, healthcare worker protection, and patient treatment. Interventions include integrating Infection Prevention and Control (IPC), surveillance, and case management functions, monthly mentorship visits, data-driven quality improvement assessments, and enhanced communication between facilities and public health authorities.

After eleven months, facility epidemic readiness scores improved from 55% to 87%. Reports of suspected reportable diseases increased from 184 to 290 per month, with 94% reported within 24 hours. A total of 75 cases of epidemic-prone diseases were detected across 17 facilities, with 99% of cases meeting the 7-1-7 target for detection, and 100% meeting the target for notification. IPC scores improved from 56% to 94%, and correct donning and doffing of PPE by HCWs improved from 34% to 87%. Bottlenecks included inconsistent supply chains and inadequate infrastructure.

ERPHC has demonstrated rapid improvements in performance, emphasising the impact of integration across technical disciplines and targeted mentorship in boosting epidemic readiness. Early results of the ERPHC approach show potential to accelerate the detection and reporting of epidemic-prone diseases and improve HCW and patient safety.

**SUMMARY BOX:**

- Primary healthcare (PHC) is critical for early outbreak detection, response, and maintaining essential health services, making it a cornerstone of health security.
- Epidemic Ready Primary Health Care (ERPHC) is a PHC system that can effectively prevent, detect and respond to outbreaks, while continuing essential health services. Achieving ERPHC requires healthcare workers (HCWs) to connect with local communities, promptly detect and report cases, protect themselves and others, and treat cases while maintaining services.
- The ERPHC pilot showed rapid improvements, with health facility performance increasing from 55% to 87% over eleven months. Seventy-five cases of epidemic-prone diseases were detected, with 99% meeting the 7-1-7 target for detection and 100% the target for notification.
- An integrated approach across technical disciplines and continuous mentorship has proven effective in accelerating HCW capacity and health facility performance.
- Strengthening supply chains and addressing infrastructure bottlenecks are crucial for HCW safety and facility epidemic readiness.

## INTRODUCTION

Epidemics begin in and are stopped by communities, placing Primary Health Care (PHC) at the forefront of epidemic preparedness and response. As the initial point of contact for most people, PHC serves as a vital link between health systems and communities. Given the critical role of PHC in delivering essential health services and community-level public health measures,^1^ robust PHC is necessary for a resilient health system.^2^ Well prepared PHC facilities can serve as the first line of defence during outbreaks, facilitating early detection and response, limiting disease transmission, and alleviating pressure on hospitals. However, PHC is often not adequately supported to fulfil this critical role, as most health security initiatives are concentrated at the national level, with PHC strengthening largely absent.^3^ This is important not only to find and stop disease threats sooner, but to maintain life-saving health services. During the 2014–2016 West African Ebola epidemic, over 10,000 additional deaths were estimated to have occurred as a result of disrupted PHC services.^4^ The consequences of this gap were also evident and deadly during the COVID-19 pandemic, with more than 90% of countries reporting disruptions to essential health services, and PHC among the most affected service delivery settings.^5^ PHC faced numerous challenges, including insufficient funding and personnel, shortages of personal protective equipment (PPE), inadequate infrastructure, limited training, lack of standardised protocols, and poor coordination with hospitals and public health authorities.^6^

This lack of preparedness undermines the ability to quickly detect, report, and safely respond to infectious diseases, a crucial aspect to contain outbreaks before they escalate.^7^ The 7-1-7 target, a framework to improve outbreak performance, assesses healthcare system timeliness to detect (≤7 days from emergence), notify public health authorities (≤1 day from detection), and complete all essential early response actions (≤7 days from notification).^8^ As outbreaks are often identified when patients promptly seek care, and astute HCWs suspect and report cases,^9^ PHC is central to achieving these targets. A multi-country analysis of 7-1-7 revealed that 61% of outbreak detection bottlenecks occurred at the health facility level, with low clinical suspicion among HCWs identified as the most frequent gap.^10^ Similarly, delays in the detection of the 2022 Uganda Ebola Sudan virus disease outbreak were attributed to low clinical suspicion among HCWs.^11^ These findings underscore the need to strengthen HCW capacity to promptly detect and report public health threats.

Beyond outbreak detection, PHC facilities without rigorous infection prevention and control (IPC) practices pose a risk for healthcare-associated transmission and HCW infection. Within the first six months of the COVID-19 pandemic, an estimated 152,888 HCWs were infected.^12^ Such infections have impacts far beyond the individual HCW. During the COVID-19 pandemic, infections among HCWs in low-and middle-income countries incurred large economic costs, with estimates ranging from $10,000–$36,000 per infection, depending on the country.^13^ These findings highlight the need to equip HCWs not only to recognise and report epidemic-prone diseases, but also to implement effective IPC measures. This manuscript offers evidence-based insights and practical recommendations to support replication and scaling of a new approach to strengthen epidemic preparedness at the PHC level.

## EPIDEMIC READY PRIMARY HEALTH CARE

Epidemic Ready Primary Health Care (ERPHC) is a PHC approach that can prevent, detect and respond to outbreaks while continuing essential health services.^9^ ERPHC connects and integrates the public health functions of epidemic preparedness and response with clinical health service delivery, ensuring PHC HCWs can quickly identify cases (speed), manage them safely (safety), and handle increases in patient volume (surge). Achieving ERPHC requires HCWs to **connect** with local communities to enhance community trust, promptly **detect** and report cases, **protect** themselves and others by applying IPC precautions, and **treat** cases while maintaining essential health services. ERPHC can aid early outbreak detection and response (achieving the 7-1-7 targets), protect patients and frontline HCWs, and ensure the continuity of essential health services during emergencies.

### EPIDEMIC READY PRIMARY HEALTH CARE PILOT

Resolve to Save Lives, in collaboration with partners and Ministries of Health (MOHs)/National Public Health institutes (NPHIs) launched the ERPHC project in December 2023. This initiative built on a previous multi-year project aimed at protecting HCWs and strengthening IPC during COVID-19,^14^ incorporating lessons learned to inform design and implementation. ERPHC has been implemented across 654 PHC facilities in Ethiopia, Nigeria, Sierra Leone, and Uganda (Table 1). The integration of three technical areas – surveillance, IPC, and case management – is fundamental to ERPHC. This approach bridges technical silos and aligns with the real-world workflows of HCWs, ensuring they acquire the necessary skills to detect and report suspected cases, protect themselves, and provide immediate patient care. The model includes continuous capacity strengthening through ongoing mentorship visits to enhance HCW knowledge, skills, and confidence. Mentors, comprising a range of local public health and healthcare workers (Table 1), were identified and trained in the ERPHC approach. Modest incentives facilitate their monthly visits to health facilities. Other interventions include systematic and periodic assessment of facility performance, combined with data-driven action planning to inform improvements as well as strengthen public health emergency management strategies at the health facility and subnational level to ensure the continuity of essential health services during health threats. Additionally, the model focuses on improving bidirectional communication and coordination between PHC health facilities and public health authorities at the subnational level to facilitate rapid detection, notification, and a coordinated response. The pilot was designed to be an adaptable, contextualised model that can be scaled within implementing countries and replicated in others.

**Table 1:** ERPHC Pilot Summary.

| **Model** | **Ethiopia** | **Nigeria** | **Sierra Leone** | **Uganda** |
| --- | --- | --- | --- | --- |
| Location | Addis Ababa and Sidama region | Kano state and Ogun state | Kenema district, Bombali district, Port Loko district, Western Area Urban and Western Area Rural districts | Kampala Metropolitan Area, West Nile region and Eastern region |
| Number of Health Facilities | 52 | 192 | 80 | 330 |
| Types of Health Facilities | Public | Public | Public | Public including prison health facilities, private for profit, and private not for profit |
| Frequency of Mentorship Visits | Monthly | Monthly | Twice per month | Monthly |
| Mentors | Addis Ababa: Public Health Emergency (PHEM) Officers from sub-city PHEM offices<br>Sidama: Hospital PHEM officers | Government public health workers from Local Government Authority and State levels | Implementing partner staff | Government healthcare workers |
| Mentor to Health Facility Ratio | 1:2–3 | 1:3–4 | 1:10 | 1:2–3 |

#### Monitoring Pilot Progress

Health facility epidemic readiness was assessed using a standardised checklist completed during mentorship visits (Supplemental Material 1). The checklist includes 23 indicators across three key domains: health facility preparedness, IPC, and surveillance. Knowledge-based indicators were assessed by randomly selecting HCWs at each facility to respond to the question. Data were collected monthly at the facility level using electronic systems such as ODK and DHIS2, with descriptive analysis performed in Excel. An overall ERPHC composite score, calculated as the mean of scores from the three domains, provided a measure of overall epidemic readiness (Table 2). In addition, an outbreak assessment tool (Supplemental Material 2) was utilized to capture timeliness data, measuring the speed of detection and notification following confirmed cases of priority pathogens at ERPHC-supported health facilities or during declared outbreaks. This tool was adapted from the 7-1-7 assessment tool, tailored for the PHC level, to assess contributions to the first “7” and the “1” of the 7-1-7 framework, and identify health facility-level bottlenecks and enablers. Insights described below are based on data trends observed during the first 11 months of implementation, up to October 2024, aggregated across the four countries.

**Table 2:**
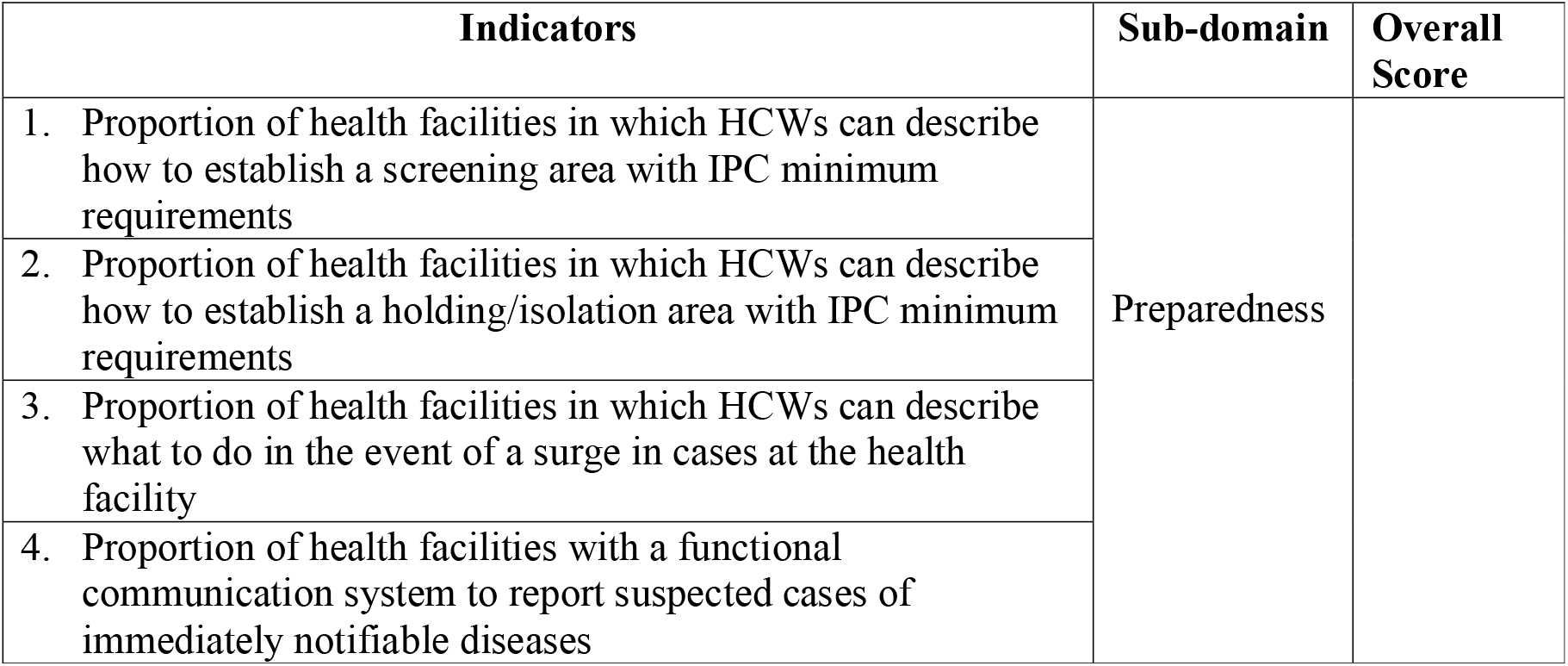
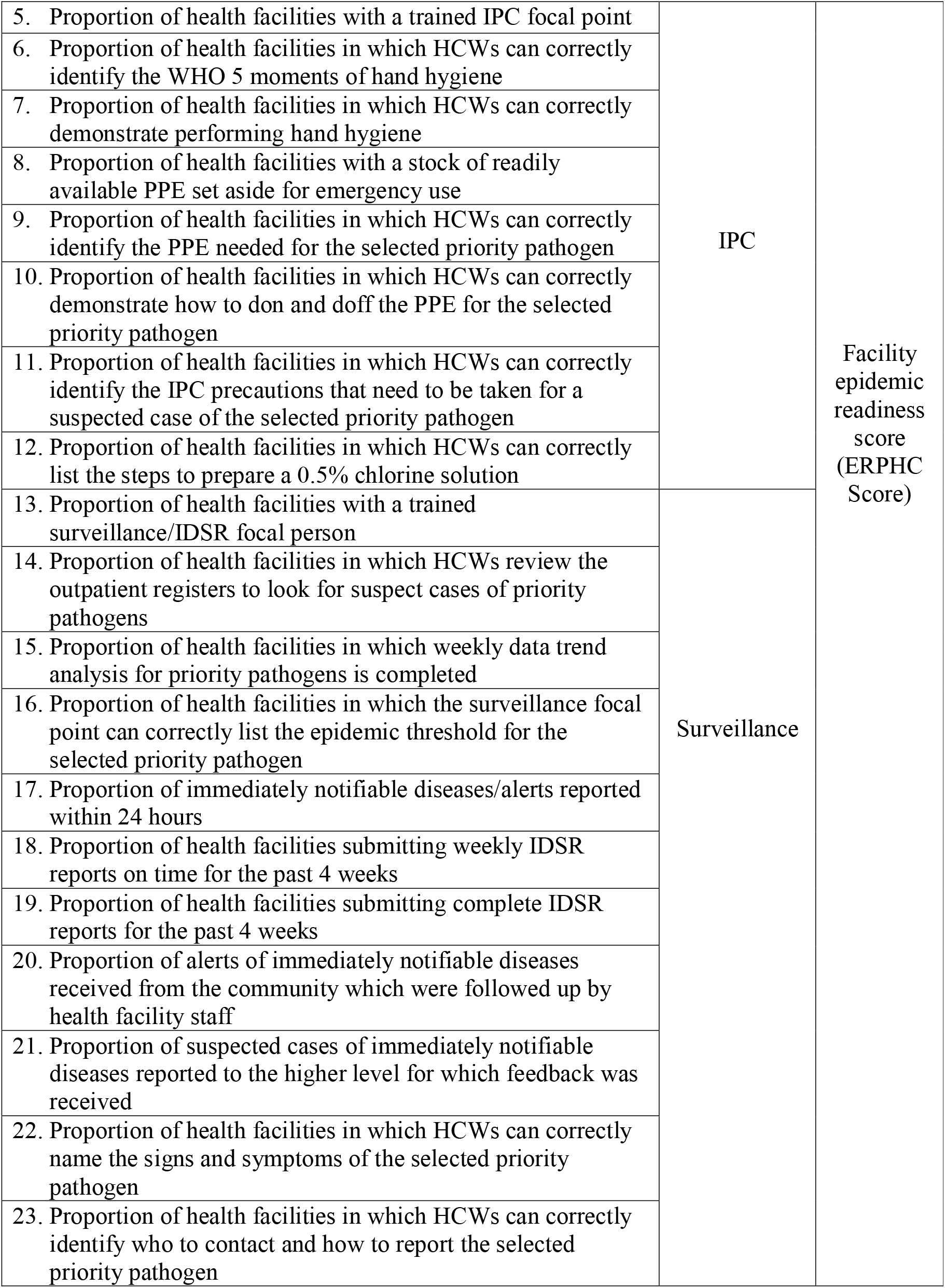
List of indicators assessed per domain comprising overall ERPHC score.

### EARLY OUTCOMES OF THE PILOT MODEL SHOW RAPID IMPROVEMENTS IN OVERALL READINESS

The pilot has demonstrated the capacity for rapid improvement of PHC epidemic readiness. Over eleven months of implementation, the average composite ERPHC score for health facilities increased from 55% at baseline to 87%, with improvement observed across all three sub-domains, across all countries (Figure 1).

**Figure 1:**
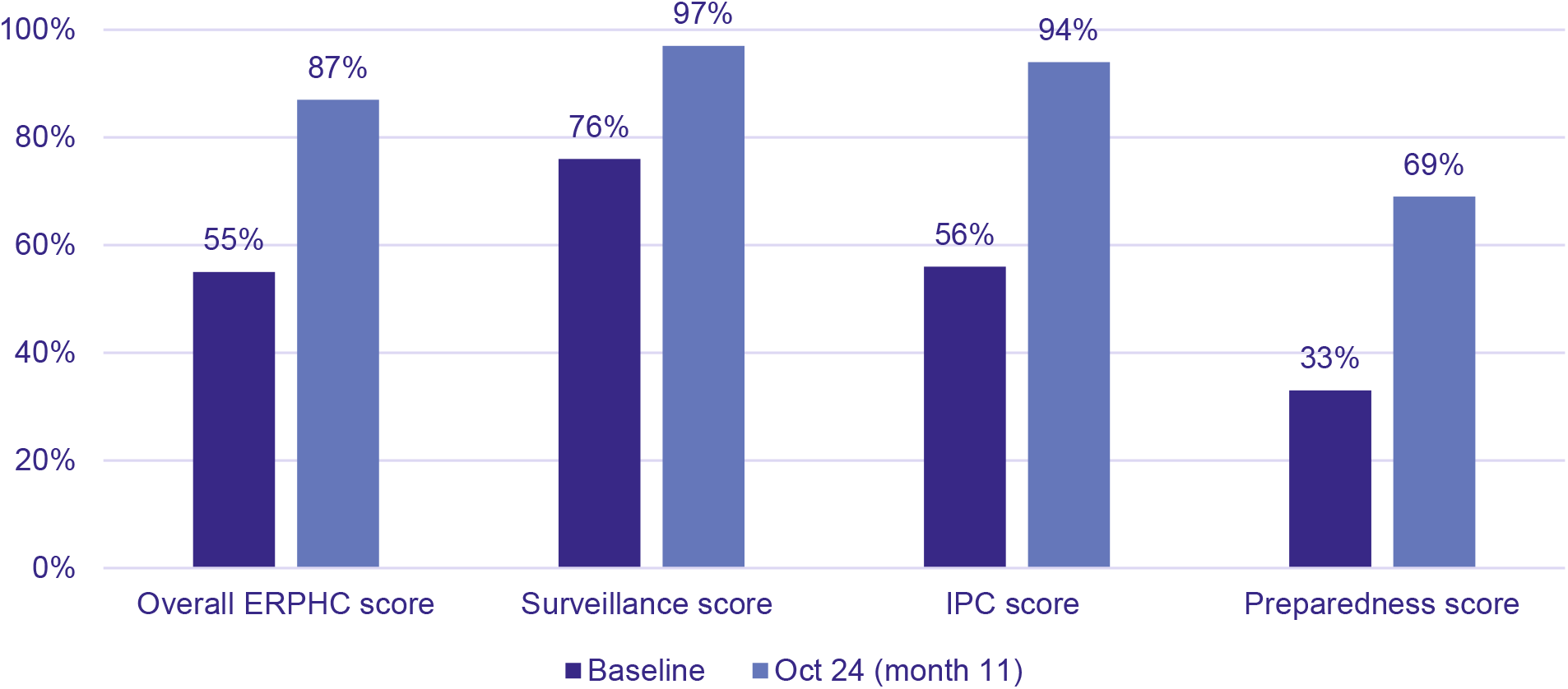
Average Health Facility ERPHC and Domain Scores (Baseline vs. October 2024)

#### Improved surveillance data analysis and reporting

The proportion of health facilities performing weekly review of outpatient registers to identify suspect cases of immediately notifiable diseases (active case searches), rose from 74% to 97%. Whilst the proportion of health facilities conducting weekly trend data analysis for priority diseases increased from 56% to 93%, and HCW knowledge of epidemic thresholds improved from 58% to 96%. Gains were also made in surveillance reporting – total monthly alerts reported from facilities increased from 184 to 290, with timely reporting (within 24 hours) rising from 81% to 94%. Feedback to the health facility following the submission of alert reports improved from 63% to 88%. The proportion of immediately notifiable alerts from the community which were followed up by health facilities increased from 82% to 100%. Additionally, the submission of complete and timely Integrated Disease Surveillance and Response (IDSR) reports improved from 86% and 85% to 98% and 97%, respectively.

#### Narrowing the gap between infection prevention and control and surveillance

At baseline, health facility IPC performance lagged behind surveillance, with scores of 56% and 76%, respectively. Eleven months into the pilot, this gap has almost closed, with IPC performance reaching 94% and surveillance performance at 97%. Substantial gains were observed in hand hygiene practices, with the proportion of facilities with HCWs able to correctly identify the WHO five moments of hand hygiene increasing from 43% to 98%, and those able to correctly perform hand hygiene rising from 47% to 98%. Availability of PPE for emergency use also improved, with 85% of facilities maintaining a stock, up from 59% at baseline. The proportion of facilities where HCWs know the requirements to establish screening and isolation areas with minimum IPC requirements increased from 20% and 17% at baseline to 66% and 58%, respectively.

#### Reducing knowledge and behaviour gaps among healthcare workers

Baseline data revealed gaps in HCW knowledge, particularly regarding the signs and symptoms of priority pathogens and the process for notifying them, compared to knowledge of the necessary IPC precautions. These gaps have largely closed, with current scores for these key indicators reaching similar levels, demonstrating progress in aligning knowledge across detection, notification, and protection (Figure 2).

**Figure 2:**
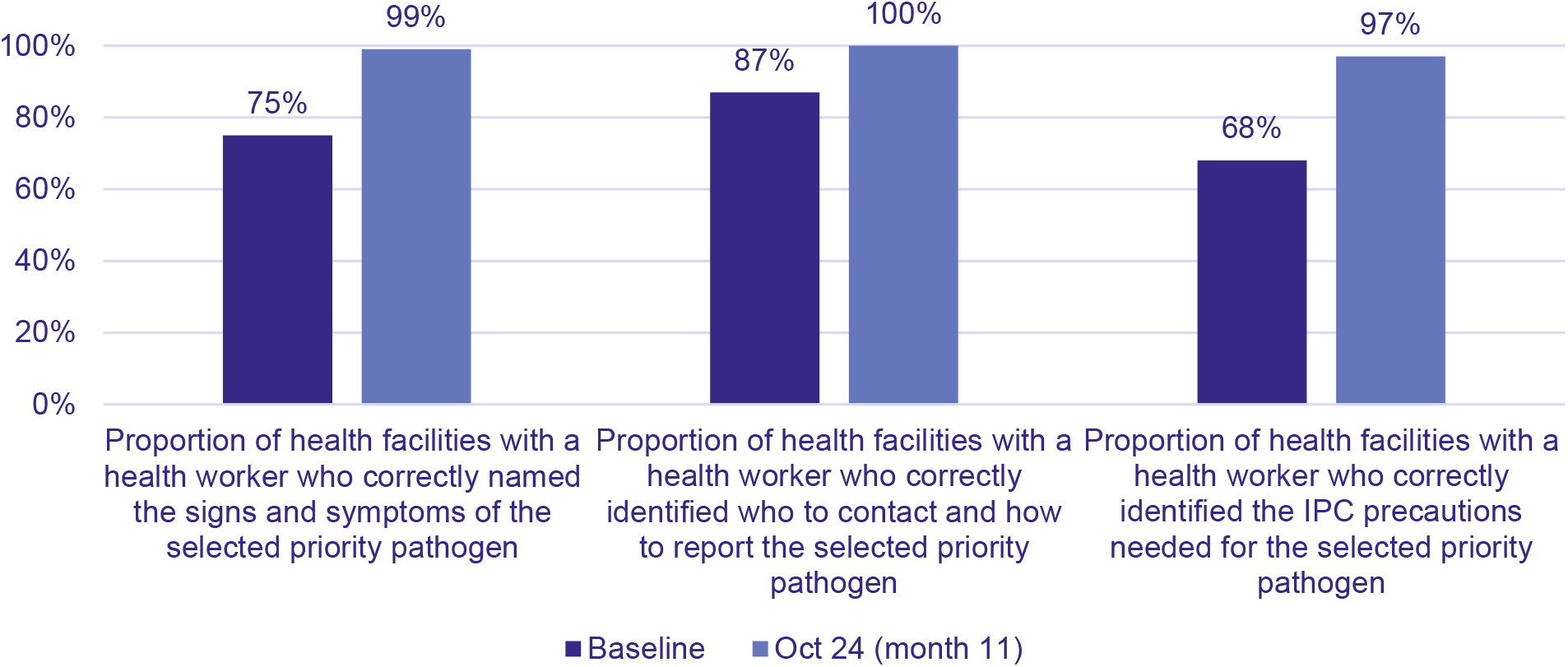
Average Health Facility Scores Across Key Knowledge Indicators (Detect, Notify, Protect) (Baseline vs. October 2024)

While HCW knowledge has improved, gaps between knowledge and practice remain. At baseline, only 34% of facilities had a HCW who could correctly demonstrate PPE donning and doffing, compared to 66% who could identify the correct PPE to wear. Whilst improved, this gap persists with current figures of 87% and 99% respectively (Figure 3).

**Figure 3:**
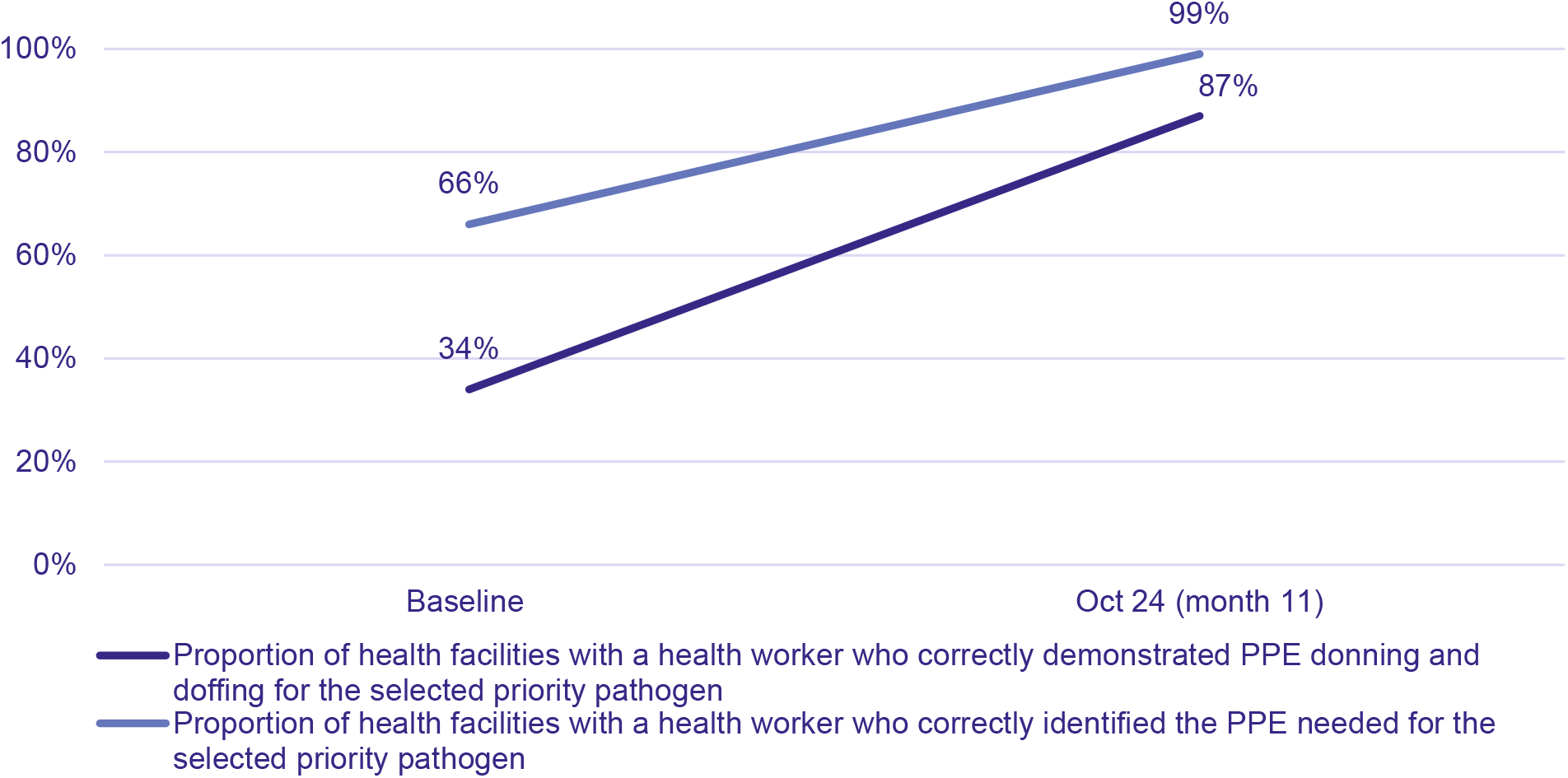
Average Health Facility Scores for PPE Knowledge and Demonstration (Baseline vs. October 2024)

### Detection and notification improvements for epidemic-prone diseases

During the implementation period, a total of 75 cases of epidemic-prone diseases were detected across 17 facilities (1 Yellow Fever, 2 Diphtheria, 3 Mpox, 4 Cholera, and 65 Measles cases). Of the 75 cases, 99% (n=74) met the 7-1-7 target for detection, and 100% met the 7-1-7 target for notification. The top three enablers supporting detection and notification included HCW knowledge of case definition (32%), reliable systems for notification (29%), and HCW knowledge of the notification process (21%).

### LESSONS LEARNED AND RECOMMENDATIONS

Initial data from the ERPHC pilot suggests that epidemic-readiness at the PHC level can be strengthened through interventions targeting HCW knowledge, skills, and behaviour.

#### Bridging technical silos to strengthen HCW capacity

Adopting an integrated approach across technical disciplines is critical to avoid fragmented training efforts and align capacity-strengthening initiatives with the real-world workflows of HCWs. The ERPHC program addresses this by integrating the technical disciplines of surveillance, IPC, and case management, ensuring HCWs are simultaneously equipped to detect and report pathogens, protect themselves, and provide immediate care for patients. Bridging technical silos empowers HCWs with the comprehensive skills needed to detect, protect, and treat, ultimately improving patient outcomes and HCW safety. However, implementing such an approach presents challenges due to traditionally siloed structures, where different technical disciplines are managed by separate departments within MOHs and NPHIs. Overcoming these silos requires coordination and collaboration to harmonise training, reporting, and response protocols, ensuring that HCWs develop skills across all relevant technical areas. Mentorship can play a critical role in bridging these challenges by providing ongoing support that helps HCWs apply and integrate skills across different disciplines, ensuring consistent adherence to best practices.

#### Mentorship is a highly adaptable approach that can drive rapid improvement

Mentorship has been shown to improve quality of care outcomes in low- and middle-income countries,^15^ strengthen health systems,^16^ and improve the clinical management of infectious diseases.^17^ In the ERPHC pilot, mentorship accelerated improvements in health facility performance, with the choice of mentors playing an important role. For instance, Nigeria exhibited quick gains in facility performance which could be attributable to their strategic choice of mentors who are full-time public health staff within the State and Local Government Authority. By leveraging their knowledge of the system, they have been able to address operational bottlenecks and mobilise relevant resources to address identified gaps. Tailoring mentorship to address both technical and operational challenges has proven essential for enhancing performance. Additionally, mentorship played a crucial role in not only strengthening the technical capacity of HCWs but also bridging the gap between theoretical knowledge and practical application. Unlike one-time training sessions, ongoing mentorship provides continuous support, promoting behaviour change, as HCWs progressively develop and apply skills in their daily practice. Furthermore, the mentorship model has proven effective across all four countries and is adaptable to various settings, including private and public health facilities, prison health services, and both urban and rural areas, demonstrating its ability to meet the needs of diverse healthcare contexts. Building on the success of mentorship in enhancing HCW capacity and health facility performance, mentorship should be more widely integrated into health systems.

#### Enablers for epidemic readiness at health facilities

Across countries, several key enablers have emerged. Strong government engagement and support, from national to sub-national levels, has been critical. This buy-in, fostered through close collaboration and initial co-design to contextualise the ERPHC model, has helped drive the project’s success. Active engagement from health facility leadership has been crucial, with stronger involvement leading to more effective implementation. Utilising communication channels such as WhatsApp facilitated ongoing mentorship and enhanced bi-directional information sharing between sub-national and facility levels. Lastly, the functionality of IDSR reporting systems have been an important enabler. Where these systems have been effective and functional for HCWs, immediate reporting of suspected alerts within 24 hours has been consistently high.

#### Bottlenecks for epidemic readiness at health facilities

Despite rapid gains, several bottlenecks have been identified as challenges to achieving epidemic readiness at health facilities. Among these, supply chain issues were particularly widespread, with inconsistent supplies of essential items including PPE, exposing HCWs to risks. Strengthening supply chains is crucial to ensure consistent availability of these materials, as these gaps not only compromise the safety and preparedness of health facilities but also the effectiveness of mentorship efforts. Physical infrastructure was another significant bottleneck, affecting capacities for screening, triage, and isolation. Many facilities lack adequate spaces for screening and triage, and isolation areas are often inadequate and poorly equipped, hindering the safe management of infectious cases. Poor Water, Sanitation, and Hygiene (WASH) infrastructure, such as reliable access to clean water, functional sanitation facilities, and proper waste disposal systems, further compromises IPC practices, putting both patients and HCWs at risk. Prioritising physical infrastructure upgrades is essential to improve facility safety and epidemic readiness.

Other bottlenecks included staff attrition, where HCW turnover disrupted project continuity. In addition, delayed feedback to health facilities following submission of surveillance alerts was a particular challenge with negative laboratory results; feedback to the facility was often delayed or neglected. These delays can undermine the effectiveness of surveillance efforts, potentially undermining the trust and motivation of HCWs to continue to report alerts.

Strengthening feedback loops is crucial to maintain consistent communication between health facilities and public health systems, ensuring timely notifications critical for early outbreak detection.

There are limitations to our findings. While the pilot spans diverse settings, the results may not be generalisable without further adaptations for new contexts. Secondly, data on individual HCW knowledge may not fully reflect overall health facility performance, particularly in larger facilities, as only one HCW was assessed and observed per visit. Lastly, some health facilities were part of a prior initiative to strengthen IPC in the context of COVID-19,^14^ and thus their performance may not be reflective of PHC facilities more broadly. Despite these limitations, the observed improvements in IPC, surveillance, and preparedness across diverse settings highlights the ERPHC model’s potential for strengthening epidemic readiness in PHC facilities.

## CONCLUSION

The ERPHC approach has demonstrated rapid improvements in the detection and reporting of epidemic-prone diseases and in IPC at PHC facilities across diverse settings. Early results underscore the effectiveness of integrated technical approaches and targeted, mentored interventions to enhance epidemic readiness at the PHC level. Scaling the ERPHC model could improve early outbreak detection, support the achievement of the 7-1-7 targets, and enhance HCW safety. Strengthening epidemic readiness at the PHC level would contribute to a more resilient health system prepared for and capable of responding to emerging health threats.

## Supporting information

Supplemental Material 1_Health Facility Epidemic Readiness Checklist

Supplemental Material 2_ Health Facility Outbreak Assessment Tool

## Acknowledgements

We thank our implementing partners and respective Ministries of Health and National Public Health Institutes in Ethiopia, Nigeria, Sierra Leone, and Uganda for their efforts in implementing this initiative. Our implementing partners include African Field Epidemiology Network (AFENET), ICAP at Columbia University, Infectious Diseases Institute at Makerere University, and Resolve to Save Lives.

## Contributors

SM conceived the paper and served as the primary author. ABC, SK, JL, LP, and AM contributed to the conceptualisation and provided substantial editorial input. MK, AMH, SMS, AW contributed data to the manuscript. SK analysed the data. All authors reviewed and edited the final draft of the manuscript. All authors were involved in the design and implementation of the ERPHC pilot.

## Ethics statements

This work was determined to be exempt human subjects research by the Resolve to Save Lives Research Committee. Approvals to disseminate findings or IRB review and approval was obtained as required in each country.

### Patient consent for publication

Not applicable.

### Provenance and peer review

Not commissioned, externally peer reviewed.

## Data availability statement

All data relevant to the study are included in the article or uploaded as online supplemental information.

