## Supplemental Material 1_Health Facility Epidemic Readiness Checklist for "Progress in Epidemic Ready Primary Health Care: Early Pilot Results from Four African Countries (Ethiopia, Nigeria, Sierra Leone and Uganda), December 2023 – October 2024"

| **Epidemic Ready Primary Health Care**<br>**Health Facility Epidemic Readiness Checklist** | | | |
|---|---|---|---|
| **Health Facility Name:** | | | |
| **Type of Primary Health Center *(i.e., HF I, II, III, IV)*:** | | | |
| **District/Province:** | | | |
| **Date of Visit:** | | | |
| **Name of person(s) using the mentorship tool:** | | | |
| **Primary Contact(s) at PHC (e.g., IPC focal point, IDSR focal point or health facility surveillance focal person, etc.):** | | | |
| **Health Care Facility Preparedness** | | **Response** | **Mentor's Notes** |
| **1** | Can a screening station with IPC minimum requirements be rapidly established at the health facility?<br><br>*Randomly select 1 HCW at time of visit and ask how he/she would set-up a screening station at the HF, ensuring that the IPC minimum requirements are incorporated in the response (see Mentorship Guide items to be incorporated into response). Each month, try to ensure a different HCW is selected.* | ○ Yes (all 9 requirements of the question are answered correctly)<br>○ Mostly: most (between 5–8) of the requirements are included in the response<br>○ Partially: some (between 1–4) of the requirements are included in the response<br>○ No (none of the requirements of the question are answered correctly) | |
| **2** | Can an isolation area/holding area with IPC minimum requirements be rapidly established at the health facility?<br><br>*Randomly select 1 HCW at time of visit and ask how he/she would establish an isolation/holding area, ensuring that the IPC minimum requirements are incorporated in the response (see Mentorship Guide on items to be incorporated into response).* | ○ Yes (all 10 requirements of the question are answered correctly)<br>○ Mostly: most (between 5–9) of the requirements are included in the response<br>○ Partially: some (between 1–4) of the requirements are included in the response<br>○ No (none of the requirements of the question are answered correctly) | |
| **3** | Can a member of the health facility staff describe what to do in case of a surge in cases at the health facility?<br><br>*Randomly select 1 health facility staff (including the leadership/in-charge) at time of visit and ask what to do in case of a surge in cases at the HF? (See Mentorship Guide on items to be incorporated into response).* | ○ Yes (all 10 requirements of the question are answered correctly)<br>○ Mostly: most (between 5–9) of the requirements are included in the response | |

| | | | |
|---|---|---|---|
| | | ○ Partially: some (between 1–4) of the requirements are included in the response<br>○ No (none of the requirements of the question are answered correctly) | |
| **4** | Does the HF have a functional communication system (i.e., airtime, data, mobile phone (either dedicated phone at HF or health staff's personal phone)) to report immediately reportable pathogens/diseases/events? | ○ Yes<br>○ No | |
| **IPC Performance** | | **Response** | **Mentor's Notes** |
| **5** | Is there an IPC focal person/point at the health facility that has received IPC training in the last 12 months? | ○ Yes<br>○ No | |
| **6** | Are health staff able to correctly identify the 5 Moments of Hand Hygiene?<br><br>*Randomly select 1 health facility staff member at time of visit and ask about the 5 Moments of Hand Hygiene. Each month, try to ensure a different HCW is selected.* | ○ Yes (all five moments correctly identified)<br>○ No (none or some of the five moments correctly identified) | |
| **7** | Are health staff able to correctly perform hand hygiene?<br><br>*Randomly select 1 health facility staff member and ask them to demonstrate correct hand hygiene (either using soap/water or ABHR).* | ○ Yes (all hand hygiene steps performed correctly)<br>○ No (none or some hand hygiene steps performed correctly) | |
| **8** | Does the health facility have a stock of readily available PPE set aside for emergency use?<br>• Gowns<br>• Gloves<br>• Surgical masks<br>• Eye protection<br><br>*Only mark yes if the health facility has* ***ALL*** *the required PPE (gowns, gloves, surgical masks, eye protection) readily available and set aside for emergency use.* | ○ Yes<br>○ No | |
| **9** | Are health staff able to correctly identify PPE needed for a select priority pathogen?<br><br>Enter name of selected priority pathogen for monthly visit (**use the same priority pathogen throughout mentorship tool**). Next month, select a different pathogen.<br><br>*Randomly select 1 HCW at time of visit and ask him/her to identify the correct PPE needed for the selected priority pathogen (see Mentorship Guide on items to be incorporated into response).* | ○ Yes<br>○ No<br><br>Insert pathogen name | |

| | | | |
|---|---|---|---|
| | *Only mark yes if **ALL** required PPE items can be identified for the selected priority pathogen* | | |
| **10a** | Are health staff able to correctly demonstrate how to don and doff PPE items for a select priority pathogen?<br><br>Enter name of selected priority pathogen for monthly visit<br><br>Next month, select a different priority pathogen.<br><br>*Randomly select 1 HCW and ask him/her to demonstrate the correct donning/doffing processes for the PPE for the selected priority pathogen. **Use the same priority pathogen throughout mentorship tool**) (see Mentorship Guide on items to be incorporated into response).* | ○ Yes<br>○ No<br><br>Insert pathogen name | |
| **10b** | Does the health facility have sufficient PPE to demonstrate how to don and doff PPE items for a select priority pathogen? | ○ Yes<br>○ No | |
| **11** | Can health staff correctly identify the IPC precautions that need to be taken for a suspected case of a selected priority pathogen?<br><br>Enter name of selected priority pathogen for monthly visit<br><br>Next month, select a different priority pathogen.<br><br>*Randomly select 1 HCW and ask him/her if they know the IPC precautions to take for a suspected case of the selected priority pathogen. **Use the same priority pathogen throughout mentorship tool** (see Mentorship Guide on items to be incorporated into response).* | ○ Yes<br>○ No<br><br>Insert pathogen name | |
| **12** | Are health staff able to correctly list steps to prepare a 0.5% chlorine solution at the health facility?<br><br>*Randomly select 1 HCW and ask him/her to list the steps for preparing a 0.5% chlorine solution for disinfection (see Mentorship Guide on items to be incorporated into response).* | ○ Yes<br>○ No | |
| **IDSR Performance** | | **Response** | **Mentor's Notes** |
| **13** | Is there an IDSR focal point, or designated person responsible for surveillance, at the HF who was trained on the latest IDSR guidelines within the last 12 months? | ○ Yes<br>○ No | |
| **14** | Has the IDSR focal point, or designate, reviewed the OPD registers (e.g., over 5 & under 5) to look for suspect cases of immediately reportable diseases/pathogens/events within the past epidemiological week (Monday–Sunday)? | ○ Yes<br>○ No | |
| **15** | Has the IDSR focal point, or designate, analysed the data from the past week to assess risk and trends of priority pathogens? | ○ Yes<br>○ No | |
| **16** | Can the IDSR focal point, or designate, correctly list the epidemic threshold for the selected priority pathogen (i.e., the number of suspect cases of a disease in a population that trigger immediate notification and response)? | ○ Yes<br>○ No | |

| | | |
|---|---|---|
| | Enter name of selected priority pathogen for monthly visit. Next month, select a different pathogen.<br><br>*Ask the focal point to share the epidemic threshold for the selected priority pathogen.* ***Use the same priority pathogen throughout mentorship tool*** *(See Mentorship Guide on items to be incorporated into response.)* | Insert pathogen name |
| **17a** | Were there any suspect cases of immediately notifiable diseases/pathogens/events within the health facility in the past week?<br><br>*Note that the answer to question 16 is based on a data review conducted by the mentor during the visit. It is up to the mentor to review the past week of OPD register data – and the IDSR weekly report for the same period – to look for immediately notifiable diseases and discuss any potential discrepancies with the IDSR focal point.* | ○ Yes<br>○ No |
| **17b** | If yes, how many? | Insert number |
| **17c** | Of those suspect cases of immediately notifiable diseases/pathogens/events, what is the number of cases that were reported to a higher level within 24 hours of the patient presenting at the health facility within the past week? | Insert number |
| **18** | Were the last 4 weekly IDSR reports sent on time to the next higher level?<br><br>*Only mark yes if* ***ALL*** *four reports were submitted on time* | ○ Yes<br>○ No |
| **19** | Were the last 4 weekly IDSR reports complete?<br><br>*Only mark yes if* ***ALL*** *four reports were complete* | ○ Yes<br>○ No |
| **20a** | Did HF staff receive any immediately notifiable alerts from the community level in the past one month? | ○ Yes<br>○ No |
| **20b** | If yes, how many? | Insert number |
| **20c** | And how many of those immediately notifiable alerts from the community level were followed up on by HF staff in the past one month? | Insert number |
| **21a** | What was the total number of suspected cases of imme-diately notifiable diseases/pathogens/events reported to the higher level, within the past one month? | Insert number |
| **21b** | What is the number of suspected cases of immediately notifiable diseases/pathogens/events reported to the higher level, within the past month, for which the health facility received feedback from the higher level? | Insert number |
| **22** | Can a healthcare worker correctly name the signs and symptoms (part of the standard case definition) of the se-lected priority pathogen?<br><br>*Randomly select 1 HCW and ask him/her to name the signs and symptoms of the selected priority pathogen (see Guidance for Mentors section on items to be incorporated into response). Each month, try to ensure a different HCW is selected.* | Partner to provide list + guidance to mentors on priority pathogens<br><br>○ Yes<br>○ No |

| | | |
|---|---|---|
| | *Only mark Yes if the HCW knows **ALL** the signs and symptoms of the selected priority pathogen.*<br><br>Enter name of selected priority pathogen for monthly visit.<br><br>**Mentors should use the same priority pathogen throughout mentorship tool** (but should not use same one each month); verify how they are diagnosed (signs and symptoms that are part of the standard case definition) and how they are recorded in the register. | Insert pathogen name |
| **23** | Can health staff correctly identify who to contact and how to report the selected immediately notifiable priority pathogen?<br><br>*Randomly select 1 HCW and ask him/her if they know who to contact and how to report the selected priority pathogen. See Mentorship Guide on items to be incorporated into response.*<br><br>*Only mark Yes if the HCW knows who to contact **AND** how to report the selected priority pathogen.* | ○ Yes<br>○ No |
