## Supplemental Material 2_ Health Facility Outbreak Assessment Tool for "Progress in Epidemic Ready Primary Health Care: Early Pilot Results from Four African Countries (Ethiopia, Nigeria, Sierra Leone and Uganda), December 2023 – October 2024"

### Epidemic Ready Primary Health Care
### Health Facility Outbreak Assessment Tool

#### Background

| | |
|---|---|
| **Pathogen** | |
| **Was an ERPHC supported health facility involved in the outbreak detection/notification?** | **Yes/No** |
| | **HF Name** |
| **Location of health facility** *(Country, District/LGA)* | |
| **Is the case confirmed by a laboratory test or Epi-link?** *(if 'No' do not continue, if 'yes' specify lab or epi-linked)* | **Yes/No** |
| **Number of cases** | |
| **Date ERPHC Outbreak Assessment Tool completed** | |

#### Section 1: Detection and notification timeliness (Speed)

This section focuses on the timeliness to detection and notification at the health facility level for each case/s. It involves capturing relevant data points to determine whether or not the health facility attained the first two targets of the 7-1-7 metric. Documenting the bottlenecks and enablers that prevent or facilitate timely action is critical for identifying best practices as well as the specific systems or processes that require strengthening.

##### Detection information

| Milestones | Date<br>DD/MM/YY | Narrative<br>(Briefly describe any key observations) |
|---|---|---|
| **Date of onset (Do)**<br>The date the patient reports they first experienced symptoms associated with the disease. *(see HF patient medical records)* | | |
| **Date of detection (Dd)**<br>Date the case was first recorded as a suspected case. This could be the date first seen at the health facility if the case was suspected. If the case was seen multiple times at the health facility, the date the case was suspected should be taken as the date of detection *(see HF registers)* | | |

##### Detection timeliness calculation

| Calculation | Target (days) | Timeliness to detection (days) (Dd-Do) | Only complete if >1 case | | Target met (Yes/No) | Narrative (Briefly describe any key observations) |
|---|---|---|---|---|---|---|
| | | | Median timeliness (days) | Range of timeliness (days) | | |
| **Timeless to detection**<br>Difference between date of onset and date of detection<br>(Dd – Do) | 7 | | | | | |

##### Notification information

| Milestones | Date<br>DD/MM/YY | Narrative<br>Briefly describe means of notification (e.g. phone, SMS) and other key observations |
|---|---|---|
| **Date of notification (Dn)**<br>Date the event is first reported by the HF to a public | | |

| |
|---|
| health authority responsible for action (see HF reports, e-IDSR systems, SMS notification, etc.) |

**Notification timeliness calculation**

| Calculation | Target (days) | Timeliness to notification (days) (Dn-Dd) | Only complete if >1 case | | Target met (Yes/No) | Narrative (Briefly describe any key observations) |
|---|---|---|---|---|---|---|
| | | | Median timeliness (days) | Range of timeliness (days) | | |
| Difference between dates of detection and notification (Dn – Dd) | 1 | | | | | |

**Bottlenecks and enablers for detection and notification at health facility level**

| Interval | Bottlenecks<br>Factors that prevented timely action. | Enablers<br>Factors that enabled timely action. |
|---|---|---|
| **Detection** | | |
| **Notification** | | |

#### Section 2: Immediate health facility case management actions (Safety)

This section should be completed as a rapid after-action clinical review with the health workers involved in managing the case. The questions focus on the essential immediate case management actions to ensure patient and health worker safety. This section should be combined with mentorship and used as a learning opportunity for health workers. If the assessment involves >1 case, only select 'yes' if the correct actions were taken for all cases. If the correct actions were taken for some and not all, select 'no' and describe further within the narrative section.

| Actions | Yes, No, NA | Narrative (Briefly describe any key observations) |
|---|---|---|
| **1) Was the case/s isolated from other patients?**<br>Discuss with Healthcare workers. This could include both use of a formal isolation/holding area if one exists or attempts to temporarily isolate the patient from others until referral. For diseases that are non-human-to-human transmission such as yellow fever and neonatal tetanus, put NA. | | |
| **2) Did the healthcare workers wear the correct PPE when managing the case/s?**<br>Correct PPE should be determined based on the mode of transmission of the pathogen. Ask health workers to recall the PPE items used. Note within the narrative section any PPE breaches or other exposures. | | |
| **3) Did the case/s receive appropriate treatment according to local SOPs, including referral for higher care if needed?**<br>Review records/registers and discuss with healthcare workers if the patient(s) received the correct treatment according to the pathogen suspected and severity of the patients' condition. In most cases this will include initial supportive care. | | |

**Bottlenecks and Enablers**

| Actions | Bottlenecks<br>Factors that prevented the correct actions. | Enablers<br>Factors that enabled the correct actions. |
|---|---|---|

| |
|---|
| **Immediate health facility case management actions** |

| Healthcare worker infections and deaths | | |
|---|---|---|
| **Were any HCWs at the HF infected?** | **Number** | **Narrative** |
| Number of HCW infections reported | | |
| Number of HCW deaths reported | | |

#### Section 3: Health facility readiness for further cases (Surge)

This section is a brief, rapid assessment of the readiness of the health facility to safely manage a surge in cases and maintain the provision of essential health services. Some of the questions within this section overlap with questions on the ERPHC mentorship tool. If this tool is being completed by the mentor during a mentorship visit, it is recommended to utilize the pathogen in question as the priority pathogen for that visit and to align the collection of some of these questions with the mentorship tool.

| Actions | Yes, No | Narrative<br>(Briefly describe any key observations) |
|---|---|---|
| **1) Are health workers at the heath facility aware of the case definition of the pathogen in question?**<br>Randomly select a health worker to answer | | |
| **2) Are health workers at the health facility aware of how and to whom to notify any further cases?**<br>Randomly select a health worker to answer | | |
| **3) Are health workers at the heath facility aware of the IPC precautions needed for the pathogen in question?**<br>Randomly select a different health worker to answer | | |
| **4) Are health workers at the heath facility aware of the clinical care/case management needed for the pathogen in question?**<br>Randomly select a health worker to answer | | |
| **5) Does the health facility have a system for screening and triage to identify further cases?**<br>Visually confirm | | |
| **6) Does the health facility have a dedicated space for a holding area/isolation area for cases if required, or a process for temporary isolation?**<br>Visually confirm | | |
| **7) Are health workers able to adapt the clinical setting to create more space for patients and/or move patients around (i.e., one-way flow of patients) the clinical setting safely during a surge?** | | |
| **8) Does the health facility have an up-to-date referral pathway for further cases?** | | |
| **9) Does the health facility have established mechanisms to engage with and rapidly disseminate information to their surrounding communities?**<br>Ask Health Facility in-charge | | |
| **10) Does the health facility have adequate supplies to manage further cases?**<br>This should include a review of the PPE items needed, as well as drugs/consumables needed for case management | | |

| **Summary and Actions**<br><br>Review the responses to sections 1–3 to complete this summary section. Results from the overall assessment should be utilized to develop an action plan for improvement with the health facility staff. | |
|---|---|
| | **Yes/No** |
| Did the health facility meet the timeliness target for detection and notification?<br>Review answers to section 1 | |
| Did the health facility complete all three of the immediate health facility actions correctly?<br>Review answers to section 2 | |
| Is the health facility ready for a potential surge in cases?<br>Review answers to section 3 (all components should be present for yes) | |

**Action plan**

| Action | Responsible | Timeline |
|---|---|---|
